# Barriers and facilitators to cervical precancer treatment adherence among women in Harare, Zimbabwe: A qualitative study

**DOI:** 10.64898/2026.11.13.26368072

**Authors:** Belinda Mutibura, Grant Murewanhema, Zvavahera Michael Chirenje, Nathan Mhungu, Bothwell Takaingofa Guzha

## Abstract

Adherence to recommended treatment after a positive cervical screening result remains suboptimal in Zimbabwe. This study explored how women interpreted a cervical precancer diagnosis and how emotional, interpersonal, economic, and health-system factors shaped treatment decisions. We conducted semi-structured in-depth interviews with 25 purposively selected women aged 25–50 years who had screened positive by visual inspection with acetic acid and were recommended for ablative or excisional treatment at two high-volume clinics in Harare. The qualitative component drew on the Health Belief Model and a phenomenological orientation to lived experience. Data were examined using combined deductive and inductive thematic analysis. Three interrelated themes were identified. First, women initially interpreted precancer through fear, shock, and uncertainty; misconceptions that precancer meant invasive cancer, infertility, or imminent death could delay action, whereas clear counselling reframed the diagnosis as treatable. Second, multilevel conditions shaped adherence. Supportive health workers, partner or family encouragement, prior awareness, and connection to HIV services facilitated treatment, while indirect costs, transport, fragmented referral pathways, scheduled excisional services, inconsistent communication, partner opposition, and sexual coercion impeded care. Third, Health Belief Model constructs operated in both directions: perceived severity motivated action when paired with credible benefits and self-efficacy, but produced avoidance when amplified by misinformation and weak support. Improving treatment completion will require standardised counselling, reliable navigation and follow-up, more accessible excisional services, and interventions that address partner dynamics and indirect costs. Screening scale-up should be matched by timely treatment capacity so that women with positive results complete the prevention pathway.

## Introduction

Cervical cancer is largely preventable, yet it remains a major source of avoidable illness and death. In 2022, an estimated 662,301 women developed cervical cancer, and 348,874 died from the disease worldwide; the burden was concentrated in countries with lower levels of human development [1]. Zimbabwe is among the highest-burden countries, with estimated age-standardised incidence and mortality rates of 68.2 and 47.9 per 100,000 women, respectively [2].

Zimbabwe introduced a national visual inspection with acetic acid (VIAC) and cervicography programme in 2012 and has progressively decentralised screening and ablative treatment [3]. The country is also expanding human papillomavirus (HPV) testing as it prepares to transition towards HPV DNA-based primary screening [4]. This transition can increase detection, but its population benefit depends on timely assessment, treatment, and follow-up after a positive result.

Loss between screening and treatment remains an important weakness in cervical cancer prevention programmes. In Harare, only 25.9% of women with a positive VIAC result received treatment at the screening site during 2012–2016, indicating substantial reliance on referral or delayed treatment pathways [3]. Across sub-Saharan Africa, limited treatment capacity, transport costs, fragmented referrals, weak follow-up systems, misinformation, and gendered barriers can interrupt the continuum from screening to treatment [5]. These challenges may be especially consequential for women living with HIV, who have a markedly increased risk of persistent HPV infection and cervical cancer [6].

Understanding adherence therefore requires attention to both service organisation and the beliefs through which women interpret a positive result. The Health Belief Model (HBM) proposes that health action is shaped by perceived susceptibility, perceived severity, perceived benefits, perceived barriers, cues to action, and self-efficacy [7,8]. Evidence across health behaviours suggests that perceived benefits, barriers, and self-efficacy are particularly important, although the model does not fully represent emotion, gendered power, or constraints outside an individual’s control [9,10].

A phenomenological orientation complements the HBM by attending to how a diagnosis is experienced and given meaning within a person’s relationships and everyday circumstances [11,12]. Combining these lenses can show why the same perceived threat motivates one woman but overwhelms another. This study therefore explored the lived experiences of women with a positive visual inspection result in Harare, the capital city of Zimbabwe and identified the emotional, behavioural, social, and health-system factors that influenced adherence to recommended cervical precancer treatment.

## Materials and methods

### Study design

This study was the qualitative component of a sequential mixed-methods project examining adherence to cervical precancer treatment in Harare. The HBM provided a sensitising framework for interview design and deductive analysis. A phenomenological orientation was used to attend to participants’ lived emotional, relational, and health-system experiences; however, the analytic method was thematic analysis rather than a formal transcendental phenomenological analysis.

### Study setting

The study was conducted at Edith Opperman Clinic and Epworth Polyclinic in Harare, Zimbabwe. Both are high-volume public-sector sites providing cervical screening and treatment. Available ablative treatment included thermal ablation and, depending on site and period, cryotherapy. Providers delivered loop electrosurgical excision procedure (LEEP) on scheduled service days, depending on provider availability and caseload. Cryotherapy was not available at Epworth during the study period.

### Participants and sampling

Eligible participants were women aged 25–50 years who had a positive result on VIAC between September 2024 and March 2025 and a documented recommendation for ablative treatment or LEEP. Women with previous treatment for cervical cancer or incomplete clinic records were excluded. The parent study operationally defined adherence as receipt of recommended treatment within 30 days of the positive screen; treatment after 30 days or no treatment was classified as non-adherence [13].

Participants were drawn from the quantitative phase and purposively selected to obtain variation in adherence status and treatment pathway. Sampling was guided by information power [14], and recruitment ended when successive interviews yielded no substantively new codes relevant to the study objectives. Both women meeting the study’s operational adherence definition and women classified as non-adherent were included.

### Data collection

An experienced female social scientist trained in qualitative interviewing, with no prior relationship with the participants, conducted semi-structured in-depth interviews. The principal investigator developed the interview guide from the study objectives and HBM constructs. The guide was pilot-tested with two eligible women who were not included in the final sample, after which minor changes were made to question wording and sequence.

Interviews took place in private rooms at the clinics in Shona or English, according to participant preference, and lasted approximately 30–60 minutes. With permission, interviews were audio-recorded. The interviewer and a second experienced social scientist transcribed recordings verbatim and translated Shona interviews into English. The principal investigator compared all transcripts with the recordings for completeness and accuracy before analysis. Transcription was completed within 48–72 hours, and field notes documented context and supported reflexive interpretation.

### Data analysis and reflexivity

Data were analysed using Braun and Clarke’s six-phase approach to thematic analysis [15]. A combined deductive–inductive strategy was used: initial coding was sensitized by HBM constructs while allowing additional codes to be generated from participants’ accounts. An experienced qualitative analyst completed the initial coding in NVivo 12, and the principal investigator reviewed the coding and developing themes. Differences in interpretation were discussed and resolved through analytic dialogue. Codes and themes were compared iteratively across transcripts and then mapped to HBM constructs where appropriate.

Reflexivity was supported through field notes and regular research-team discussions. The team examined how clinical assumptions and prior expectations might shape interpretation and actively sought accounts that complicated or contradicted emerging themes. Reporting was guided by the Consolidated Criteria for Reporting Qualitative Research (COREQ) [16].

### Ethics

Ethical approval was obtained from the University of Zimbabwe Joint Research Ethics Committee (JREC/377/23) and the Medical Research Council of Zimbabwe (MRCZ/B/2750) before recruitment. Each participant provided written informed consent before the interview and simultaneously consented to audio-recording on the same form. Participant numbers rather than names are used with quotations as participant identifiers.

## Results

### Participant characteristics

Twenty-five women with a positive cervical screening result participated. Their ages ranged from 25 to 49 years (median 37.0 years, interquartile range 32.5–43.0). Most were married (18/25, 72%), had secondary education (21/25, 84%), and were self-employed (16/25, 64%); 22 (88%) were living with HIV. By the time of interview, 12 women had received LEEP and 13 had received thermal ablation (Table 1). The sample included women from both operational adherence groups.

**Table 1.** Sociodemographic and clinical characteristics of participants (N = 25)

| **Characteristic** | **Summary** |
|---|---|
| Age, median (IQR), years | 37.0 (32.5–43.0) |
| Education: primary | 3 (12%) |
| Education: secondary | 21 (84%) |
| Education: tertiary | 1 (4%) |
| Employment: self-employed | 16 (64%) |
| Employment: unemployed | 9 (36%) |
| Marital status: married | 18 (72%) |
| Marital status: divorced | 0 (0%) |
| Marital status: single | 6 (24%) |
| Marital status: widowed | 1 (4%) |
| HIV status: living with HIV | 22 (88%) |
| HIV status: HIV-negative | 3 (12%) |
| Age at sexual debut, median (IQR), years | 19.0 (17.0–20.0) |
| Age at sexual debut: <18 years | 6 (24%) |
| Age at sexual debut: 18–24 years | 19 (76%) |
| Treatment received by interview: LEEP | 12 (48%) |
| Treatment received by interview: thermal ablation | 13 (52%) |
| Treatment received by interview: cryotherapy | 0 (0%) |

### Theme 1: Emotional meaning-making after diagnosis

The first reaction to a positive result was often fear rather than a clear distinction between precancer and invasive cancer. Women described shock, anxiety, and thoughts of imminent death. Participant 10 explained:

> *“I didn’t think they were just pre-cancerous cells. I thought it was cancer. I got very stressed and didn’t know who to tell, where to start or what to do*.*” (Participant 10)*

The diagnosis was interpreted within existing life pressures. Relationship breakdown, HIV treatment, financial strain, and caregiving responsibilities intensified distress. One woman recalled:

> *“That was around the time my husband had just left, and I was on ART, so I was stressed* … *then I got diagnosed for cervical cancer. It was hard*.*” (Participant 6)*

Personal stories, community accounts, and social media images often collapsed the distinction between precancer and advanced disease. These narratives sometimes produced fatalism, fears of infertility, or fear that screening and treatment could cause cancer. Maternal responsibility, however, could transform fear into a reason to act. Participant 13 described thinking about her young children and becoming calmer after counselling.

> *“I was now thinking of my children as they are still young, so I am about to die* … *through support and counselling I became settled*.*” (Participant 13)*

Clear explanations helped women reinterpret the result as an early, treatable condition rather than a death sentence. Treatment then became associated with relief, hope, and restored control:

> *“I was happy that the pre-cancerous cells were discovered at an early stage* … *at this stage it was more treatable than getting to know it when it has reached a critical stage*.*” (Participant 12)*

### Theme 2: Interpersonal, economic, and health-system influences

#### Counselling and relationships with health workers

Counselling quality shaped whether fear became action or avoidance. Women valued explanations of treatment options, reassurance about side effects, respectful treatment, and the ability to contact a provider after leaving the clinic. Participant 11 described how information created urgency without removing hope:

*“I understood that I got there on time and if I had delayed, it might have been difficult to treat* … *So, I understood about the treatment options*.*” (Participant 11)*

In contrast, a brief notification without explanation amplified uncertainty. One participant said, “They just said I was positive but didn’t explain what it meant” (Participant 12). Some women emphasized that treatment itself was free, but clear communication and continuity varied by provider and visit.

#### Partner and family dynamics

Partners and relatives could provide reminders, transport, money, emotional reassurance, and support for the recommended period of sexual abstinence. A supportive partner called to check on one woman’s condition after treatment and thanked God that she was well. Family members also reminded participants about review dates.

Other women described partner opposition, limited autonomy, or sexual coercion. These experiences made adherence a negotiation within intimate relationships rather than an individual decision.

> *“One and a half months may be too long for a man not to have sex with you, and he didn’t understand this. He just forced me because he is my husband, and I was not supposed to say much*.*” (Participant 17)*

#### Financial and treatment-pathway barriers

Even where treatment was provided without a direct fee, transport, lost income, and competing household needs made repeated visits difficult. Participants described referral processes, long waits, delayed results, and uncertainty about when services would be available. The scheduled nature of LEEP added travel and timing demands that were less prominent for same-day ablative treatment. These accounts do not establish a comparative effect of treatment modality, but they identify pathway complexity as a practical barrier.

> *“Though it’s free, you may decide to stop because you’ll be stressed, and you are overwhelmed*.*” (Participant 1)*

Misinformation compounded structural barriers. Women reported rumours that instruments used for screening caused cancer, that treatment would remove the womb, or that ‘burning’ lesions would make cancer spread. Such messages undermined trust and sometimes made women postpone treatment until symptoms or fear became more severe.

#### HIV services and peer encouragement

For some women living with HIV, routine contact with HIV services created a pathway into cervical screening and treatment. Participant 18 explained that her precancer was detected through this ongoing care. Several women also described encouraging others to be screened and treated after their own experience, turning personal recovery into a cue to action within families and communities.

> *“I will say go to your nearest clinic* … *the process is not painful* … *I will encourage her to go before it compromises her health by giving her my example*.*” (Participant 13)*

### Theme 3: Health Belief Model pathways to action or avoidance

The six HBM constructs were identifiable across participants’ accounts, but none acted uniformly. Perceived severity prompted treatment when women also believed that treatment was effective and achievable; the same severity produced withdrawal when precancer was interpreted as inevitable death. Perceived barriers reflected both beliefs and material constraints. Cues to action included provider advice, symptoms, reminders, and the experiences of relatives or peers. Self-efficacy increased when women received clear counselling and practical support but diminished under financial strain, misinformation, or partner control (Table 2).

**Table 2.**
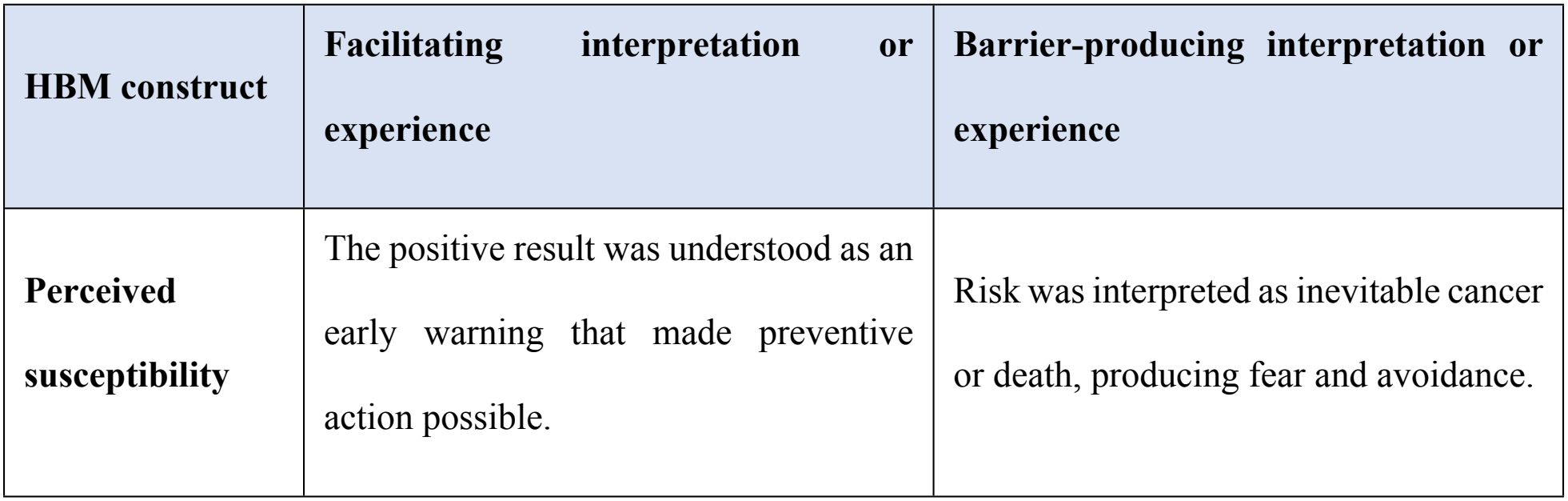
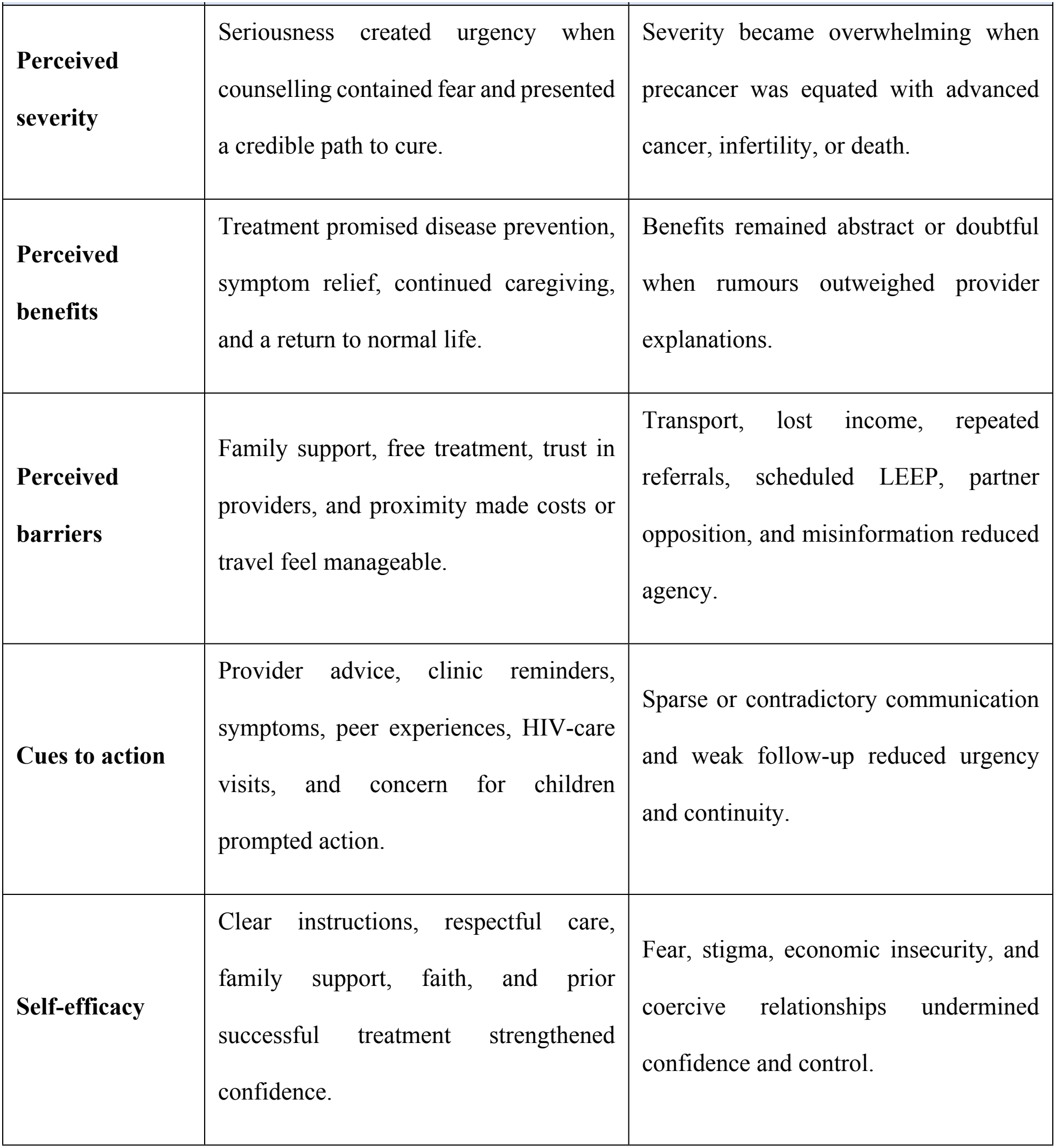
How Health Belief Model constructs facilitated or impeded treatment engagement.

| **HBM construct** | **Facilitating interpretation or experience** | **Barrier-producing interpretation or experience** |
|---|---|---|
| **Perceived susceptibility** | The positive result was understood as an early warning that made preventive action possible. | Risk was interpreted as inevitable cancer or death, producing fear and avoidance. |
| **Perceived severity** | Seriousness created urgency when counselling contained fear and presented a credible path to cure. | Severity became overwhelming when precancer was equated with advanced cancer, infertility, or death. |
| **Perceived benefits** | Treatment promised disease prevention, symptom relief, continued caregiving, and a return to normal life. | Benefits remained abstract or doubtful when rumours outweighed provider explanations. |
| **Perceived barriers** | Family support, free treatment, trust in providers, and proximity made costs or travel feel manageable. | Transport, lost income, repeated referrals, scheduled LEEP, partner opposition, and misinformation reduced agency. |
| **Cues to action** | Provider advice, clinic reminders, symptoms, peer experiences, HIV-care visits, and concern for children prompted action. | Sparse or contradictory communication and weak follow-up reduced urgency and continuity. |
| **Self-efficacy** | Clear instructions, respectful care, family support, faith, and prior successful treatment strengthened confidence. | Fear, stigma, economic insecurity, and coercive relationships undermined confidence and control. |

## Discussion

This study shows that treatment adherence after a positive cervical screening result was not a simple response to referral advice. Women first had to make sense of the diagnosis, often while navigating HIV care, caregiving responsibilities, economic insecurity, and intimate relationships. Fear was therefore neither purely a barrier nor purely a motivator. It moved women towards treatment when counselling made the condition understandable and treatable, but towards avoidance when misinformation or weak support made the threat feel uncontrollable.

Misconceptions that precancer was already invasive cancer, that treatment caused infertility, or that screening instruments caused cancer resemble barriers reported in qualitative and mixed-methods evidence from other African settings [17]. The present findings extend this literature by situating these beliefs after a positive result, when the clinical need shifts from screening uptake to treatment completion. This distinction matters because a programme may achieve screening coverage while still failing to prevent cancer if positive results are not linked to effective care.

Participants’ accounts also reflected health-system constraints previously described in Zimbabwe, including unclear referrals, limited treatment capacity, indirect costs, and discontinuity of care [18]. Similar barriers have been reported across low- and middle-income countries [19]. In this study, participants described the scheduled and multistep LEEP pathway as particularly difficult to navigate. Because the sample was qualitative and adherence-group denominators were not analysed comparatively here, these accounts should not be interpreted as proving that LEEP caused non-adherence. They instead identify service design as a plausible, modifiable influence that warrants quantitative evaluation.

Integration with HIV and sexual and reproductive health services may support continuity by using established clinic visits, records, and trusted providers [20]. This is especially relevant in a sample in which 88% of participants were living with HIV. Integration should, however, extend beyond offering a screening test: positive results require clear navigation, timely treatment appointments, and follow-up systems. Without these components, additional screening can increase the number of women waiting for care without improving treatment completion.

Partner and family relationships had a dual role. Practical and emotional support strengthened adherence, whereas opposition to treatment or post-treatment abstinence constrained women’s autonomy. Related studies have shown that partner and gender dynamics influence engagement with reproductive and cervical cancer services [21]. The report of sexual coercion in this study also indicates that counselling directed only at the woman may be insufficient. Voluntary, safety-conscious partner engagement and referral pathways for gender-based violence should be considered, while avoiding approaches that make partner permission a condition of care.

The HBM was useful for organising how women understood susceptibility, severity, benefits, barriers, cues, and self-efficacy. Its strongest contribution was to show that the effect of perceived severity depended on perceived benefits and control, consistent with broader adherence evidence [9,22]. A phenomenological orientation added the emotional and relational context that the HBM alone does not capture. Community narratives and spiritual interpretations were not peripheral; they helped constitute the meaning of the diagnosis, echoing ethnographic work on cervical cancer beliefs in East Africa [23].

These findings have direct implications for Zimbabwe’s cervical cancer elimination efforts. The World Health Organisation’s 90–70–90 strategy requires 90% of women with cervical precancer to receive treatment, not merely 70% screening coverage [24,25]. As Zimbabwe prepares to expand HPV DNA-based primary screening [4], implementation plans should match testing capacity with treatment capacity. Priority actions include standardised diagnosis-specific counselling; active patient navigation and reliable result notification; appointment reminders; monitoring of the screen-positive-to-treatment interval; and decentralisation of appropriate treatment. Programmes should also evaluate regulated task-sharing and more frequent access to LEEP where workforce, training, quality assurance, and referral safeguards permit.

This study has limitations. Participants were recruited from women identified through clinic records and were therefore already engaged with services to some extent; the experiences of women completely lost after screening may be underrepresented. Data were collected at two urban clinics, limiting transferability to rural settings and other service configurations. Interviews may have been affected by recall and social-desirability bias, particularly for sensitive relationship experiences. The study did not interview partners, healthcare workers, or programme managers, so it did not triangulate their perspectives on pathway barriers. Finally, the qualitative design identifies mechanisms and meanings but does not estimate their prevalence or causal effects.

Strengths of this study include purposive inclusion of different treatment pathways and adherence experiences, use of both deductive and inductive coding, and integration of a behavioural framework with attention to lived experience. The findings provide locally grounded targets for intervention and generate hypotheses that can be tested using the quantitative phase of the parent study.

## Conclusions

Cervical precancer treatment adherence in Harare was shaped by the interaction of diagnostic meaning, health beliefs, social relationships, economic conditions, and service organisation. Clear counselling and respectful care could transform fear into purposeful action, while misinformation, partner coercion, indirect costs, and complex referral or LEEP pathways undermined treatment completion. Zimbabwe’s expansion of cervical screening should therefore be accompanied by timely treatment capacity, patient navigation, and interventions that address both material barriers and women’s relational safety.

## Data Availability

All relevant data has been submitted in the manuscript. Any additional data shall be provided upon request from the corresponding author

## Data availability

The interview transcripts are not publicly available because they contain sensitive narratives that may permit deductive identification. A de-identified coding framework and selected de-identified data may be made available to qualified researchers upon approval by MRCZ and subject to the consent and ethics conditions governing this study. Requests should be directed to the corresponding author through the author’s email address.

## Acknowledgments

The authors thank the women who participated in the study and the clinic staff and research personnel who supported recruitment, interviewing, and data management. The authors would also want to acknowledge Dr Tapiwa Chapupu for assistance in manuscript construction and editing.

## References

1. Bray F, Laversanne M, Sung H, Ferlay J, Siegel RL, Soerjomataram I, et al. Global cancer statistics 2022: GLOBOCAN estimates of incidence and mortality worldwide for 36 cancers in 185 countries. CA Cancer J Clin. 2024;74(3):229–263. doi: 10.3322/caac.21834.

2. Ferlay J, Ervik M, Lam F, Laversanne M, Colombet M, Mery L, et al. Global Cancer Observatory: Cancer Today [Internet]. Lyon: International Agency for Research on Cancer; 2024 [cited 2026 Aug 25]. Available from: https://gco.iarc.who.int/today/

3. Gabaza C, Chonzi P, Chadambuka A, Shambira G, Juru TP, Gombe NT, et al. Utilization and outcomes of cervical cancer screening services in Harare City, 2012–2016: a secondary data analysis. BMC Health Serv Res. 2019;19:454. doi: 10.1186/s12913-019-4314-8.

4. Murewanhema G, Dzobo M, Moyo E, Moyo P, Mhizha T, Dzinamarira T. Implementing HPV-DNA screening as primary cervical cancer screening modality in Zimbabwe: challenges and recommendations. Sci Afr. 2023;21:e01889. doi: 10.1016/j.sciaf.2023.e01889.

5. Dzinamarira T, Moyo E, Dzobo M, Mbunge E, Murewanhema G. Cervical cancer in sub-Saharan Africa: an urgent call for improving accessibility and use of preventive services. Int J Gynecol Cancer. 2023;33(4):592–597. doi: 10.1136/ijgc-2022-003957.

6. Stelzle D, Tanaka LF, Lee KK, Khalil AI, Baussano I, Shah ASV, et al. Estimates of the global burden of cervical cancer associated with HIV. Lancet Glob Health. 2021;9(2):e161–e169. doi: 10.1016/S2214-109X(20)30459-9.

7. Rosenstock IM, Strecher VJ, Becker MH. Social learning theory and the Health Belief Model. Health Educ Q. 1988;15(2):175–183. doi: 10.1177/109019818801500203.

8. Champion VL, Skinner CS. The Health Belief Model. In: Glanz K, Rimer BK, Viswanath K, editors. Health behavior and health education: theory, research, and practice. 4th ed. San Francisco: Jossey-Bass; 2008. pp. 45–65.

9. Carpenter CJ. A meta-analysis of the effectiveness of Health Belief Model variables in predicting behavior. Health Commun. 2010;25(8):661–669. doi: 10.1080/10410236.2010.521906.

10. Jones CJ, Smith H, Llewellyn C. Evaluating the effectiveness of Health Belief Model interventions in improving adherence: a systematic review. Health Psychol Rev. 2014;8(3):253–269. doi: 10.1080/17437199.2013.802623.

11. Husserl E. Ideas: general introduction to pure phenomenology. London: George Allen & Unwin; 1931.

12. Moustakas C. Phenomenological research methods. Thousand Oaks: SAGE Publications; 1994.

13. Barchi F, Winter SC, Ketshogile FM, Ramogola-Masire D. Adherence to screening appointments in a cervical cancer clinic serving HIV-positive women in Botswana. BMC Public Health. 2019 Dec;19(1):318. doi:10.1186/s12889-019-6638-z,

14. Malterud K, Siersma VD, Guassora AD. Sample size in qualitative interview studies: guided by information power. Qual Health Res. 2016;26(13):1753–1760. doi: 10.1177/1049732315617444.

15. Braun V, Clarke V. Using thematic analysis in psychology. Qual Res Psychol. 2006;3(2):77–101. doi: 10.1191/1478088706qp063oa.

16. Tong A, Sainsbury P, Craig J. Consolidated criteria for reporting qualitative research (COREQ): a 32-item checklist for interviews and focus groups. Int J Qual Health Care. 2007;19(6):349–357. doi: 10.1093/intqhc/mzm042.

17. Black E, Hyslop F, Richmond R. Barriers and facilitators to uptake of cervical cancer screening among women in Uganda: a systematic review. BMC Womens Health. 2019;19:108. doi: 10.1186/s12905-019-0809-z.

18. Tapera O, Dreyer G, Kadzatsa W, Nyakabau AM, Stray-Pedersen B, Hendricks SJH. Health system constraints affecting treatment and care among women with cervical cancer in Harare, Zimbabwe. BMC Health Serv Res. 2019;19:829. doi: 10.1186/s12913-019-4697-6.

19. Chidyaonga-Maseko F, Chirwa ML, Muula AS. Underutilization of cervical cancer prevention services in low and middle income countries: a review of contributing factors. Pan Afr Med J. 2015;21:231. doi: 10.11604/pamj.2015.21.231.6350.

20. White HL, Meglioli A, Chowdhury R, Nuccio O. Integrating cervical cancer screening and preventive treatment with family planning and HIV-related services. Int J Gynaecol Obstet. 2017;138 Suppl 1:41–46. doi: 10.1002/ijgo.12194.

21. Mutyaba T, Faxelid E, Mirembe F, Weiderpass E. Influences on uptake of reproductive health services in Nsangi community of Uganda and their implications for cervical cancer screening. Reprod Health. 2007;4:4. doi: 10.1186/1742-4755-4-4.

22. DiMatteo MR, Haskard KB, Williams SL. Health beliefs, disease severity, and patient adherence: a meta-analysis. Med Care. 2007;45(6):521–528. doi: 10.1097/MLR.0b013e318032937e.

23. Gatune JW, Nyamongo IK. An ethnographic study of cervical cancer among women in rural Kenya: is there a folk causal model? Int J Gynecol Cancer. 2005;15(6):1049–1059. doi: 10.1136/ijgc-00009577-200511000-00007.

24. Canfell K, Kim JJ, Brisson M, Keane A, Simms KT, Caruana M, et al. Mortality impact of achieving WHO cervical cancer elimination targets: a comparative modelling analysis in 78 low-income and lower-middle-income countries. Lancet. 2020;395(10224):591–603. doi: 10.1016/S0140-6736(20)30157-4.

25. World Health Organization. Global strategy to accelerate the elimination of cervical cancer as a public health problem. Geneva: World Health Organization; 2020. Available from: https://www.who.int/publications/i/item/9789240014107

